# The amplified second outbreaks of global COVID-19 pandemic

**DOI:** 10.1101/2020.07.15.20154161

**Authors:** Jianping Huang, Xiaoyue Liu, Li Zhang, Kehu Yang, Yaolong Chen, Zhongwei Huang, Chuwei Liu, Xinbo Lian, Danfeng Wang

**Author notes:** Correspondence author: Jianping Huang.

## Abstract

COVID-19 is now in an epidemic phase, with a second outbreak likely to appear at any time. The intensity and timing of a second outbreak is a common concern worldwide. In this study, we made scenario projections of the potential second outbreak of COVID-19 using a statistical-epidemiology model, which considers both the impact of seasonal changes in meteorological elements and human social behaviors such as protests and city unblocking. Recent street protests in the United States and other countries are identified as a hidden trigger and amplifier of the second outbreak. The scale and intensity of subsequent COVID-19 outbreaks in the U.S. cities where the epidemic is under initial control are projected to be much greater than those of the first outbreak. For countries without reported protests, lifting the COVID-19 related restrictions prematurely would accelerate the spread of the disease and place mounting pressure on the local medical system that is already overloaded. We anticipate these projections will support public health planning and policymaking by governments and international organizations.

## 1 Introduction

Recently, the COVID-19 pandemic has spread rapidly and poses a dire threat to global public health, which claimed over 0.49 million lives, along with 9.8 million confirmed cases as of June 28^th 1^. Beyond the spread itself, the outbreak may have far-reaching consequences, negatively affecting the economic development worldwide and posing a series of long-standing social problems^2,3^. There is an urgent need for a global prediction system that can provide scientific guidelines for the World Health Organization and international decision-makers to implement effective containment measures capable of curbing the spread of COVID-19^4^. Researchers worldwide have developed various models with mathematical and statistical methods, including stochastic simulations, lognormal distribution^5^, machine learning, and artificial intelligence^6^. Among them, the susceptible-infectious-removed infectious disease model (SIR) is the most widely used^7–9^. However, this simple model is built under a series of idealized assumptions, which may limit the accuracy and reliability of the prediction. In order to obtain the prediction results with higher credibility, more complex models with fewer assumptions should be developed so as to simulate the actual situations in a more realistic manner^10^.

Although it is difficult to establish an accurate epidemiological model describing the spread of a pandemic, the reported global pandemic data contain particular solutions to the mathematical equations incorporated in epidemiological models^3,6^. It is theoretically possible to remedy the defects of prior epidemiological models by introducing the latest pandemic data and hence improve the pandemic prediction^2,4,6^. Based on this idea, we have developed a Global Prediction System of the COVID-19 Pandemic (GPCP)^11^. The system develops a modified version of the SIR model and determines the parameters through historical data fitting^12,13^, which allows it to make targeted predictions for various countries and obtain better prediction results. The first version of GPCP (CPCP-1) can capture the major features of the daily number of confirmed new cases and provides reliable predictions. However, the prediction of GPCP-1 is only valid for one month^11^.

In this study, the second version of the Global Prediction System for COVID-19 Pandemic (GPCP-2) is developed based on a modified SEIR model^14^. The system considers both the seasonal changes of meteorological elements and human social behaviors including protests and city unblocking. The paper is arranged as follows: the details of the datasets and the methodology used are given in section 2. In section 3, projections of 12 cities in the United States are presented. The projections of 15 countries with reported protests and 15 countries without reported protests are shown in section 4 and section 5, respectively. Discussion and conclusion are presented in section 6.

## 2 Method

### 2.1 The modified SEIR model

The second version of Global Prediction System for COVID-19 Pandemic (GPCP) is built based on a modified SEIR model^15^. The traditional SEIR model^10,14^ defines seven states of the disease: susceptible cases (S), insusceptible cases (P), potentially infected cases (E, infected cases in a latent period), infectious cases (I, infected cases that have not been quarantined), quarantined cases (Q, confirmed and quarantined cases), recovered cases (R), and cases of mortality (D). The SEIR model is able to emulate the time curve of an outbreak. The model is consisted of the following equations:

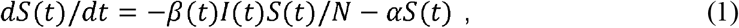

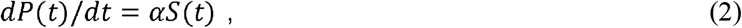

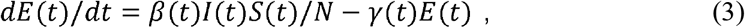

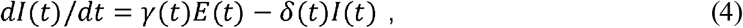

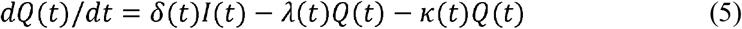

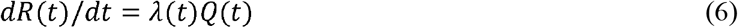

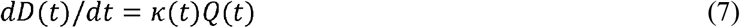

The sum of the six categories is equal to the total population (N) at any time.

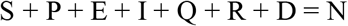

We modified the model by introducing the timing of community reopening collected from news reports. If the timing collected from news reports is not explicit enough as an input to our model, the timing will be indicated by the daily new cases on the day of reopening (dQ_c_). As the number of newly confirmed cases on a given day falls lower than dQ_c_, local authority begins to lift or loose the lockdowns.

In addition, the temporal variation of transmission rate due to changes in local temperature as well as human behaviors are considered. Generally, the transmission rate (*β*(*t*)) can be expressed by the following equations:

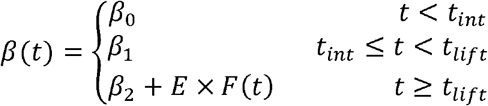

where *β*_0_ represents the transmission rate in the non-intervention period at the early stage of the pandemic (*t*<*t*_*int*_); *β*_1_ represents the transmission rate during the intervention period (*t*_*int* ≤_ *t*_*lift*_); *β*_2_ represents the transmission rate after the restriction is lifted. *β*_0_ and *β*_1_ are fitted against the actual reported data, while *β*_2_ is the assumed value in possible future scenarios. We assume a 14-day delay in the effect of the intervention on the infection rate. *F*(*t*) is the PDF function obtained by Huang et al.^16^, who found that 60.0% of confirmed COVID-19 cases occurred in places where the air temperature ranged from 5°C to 15°C. Using the NCEP reanalysis data, we calculated the global distribution of probability distribution function (PDF) values on each day of the year and included its influence on the infection rate. Figure 1 shows the PDF values for the four seasons in a year. High PDF values correspond to the ambient temperature that is conducive for the virus to spread. For the northern hemisphere, the optimal band generally moves northward in summer (June, July, and August) and moves southward in winter (December, January, and February), while for southern hemisphere the optimal band moves southward in summer (December, January, and February) and moves northward in winter (June, July, and August).

**Figure 1:**
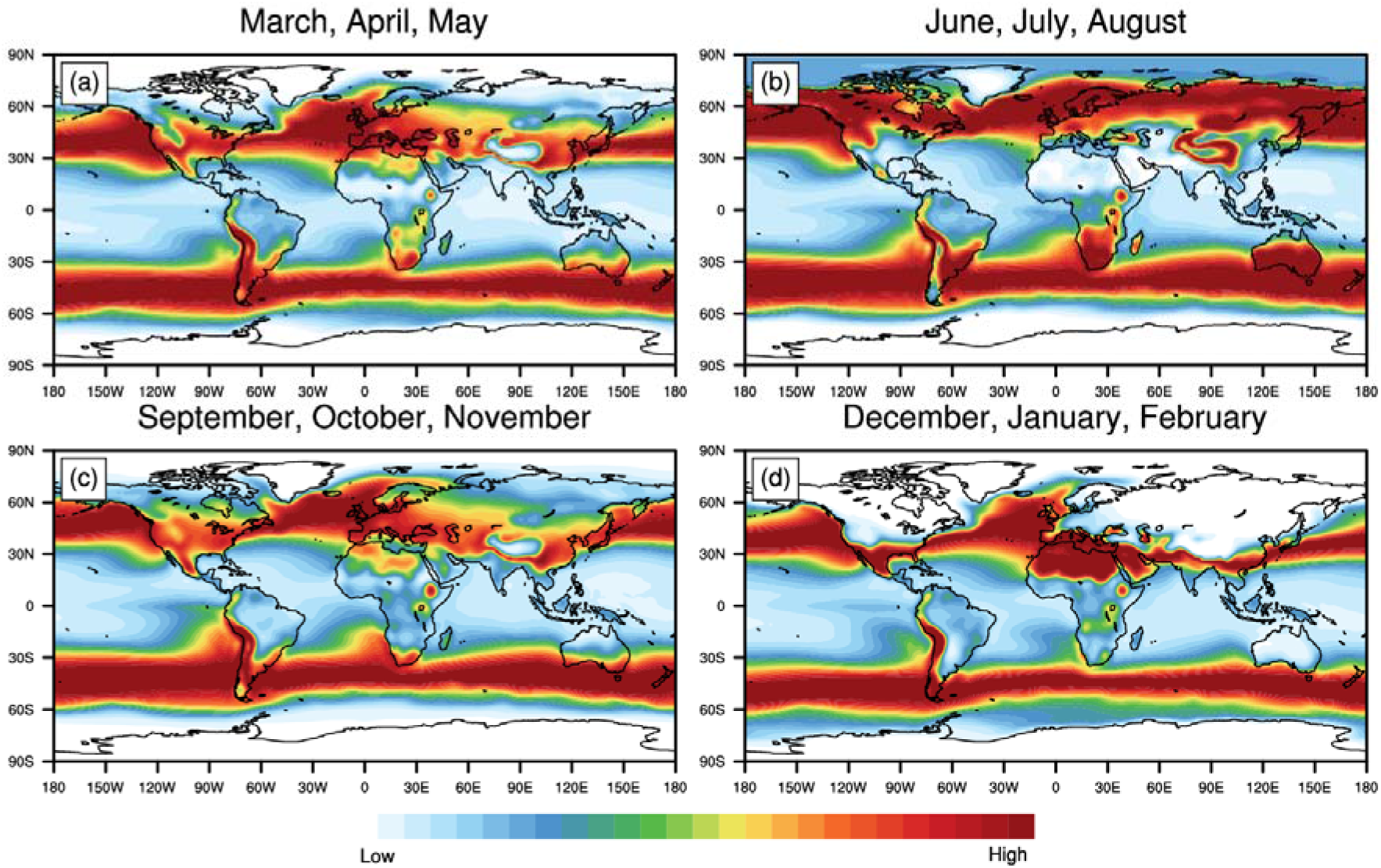
The optimal temperature zone for the spread COVID-19. Regions with warm shadings indicate more conducive temperature for the spread of the virus and vice versa.

Since the seasonality of transmission is still disputed and future trajectory of the outbreaks may be influenced by the intensity of intervention measures, four future scenarios are designed to project the epidemic after easing COVID-19 related restrictions:

- **Scenario 1**: The restrictions are completely lifted after *t*_*int*_ (*β*_2_ = *β*_0_). The seasonal forcing on the transmission rate is considered (*E* = 1).
- **Scenario 2**: The restrictions are partially lifted after *t*_*int*_ (*β*_2_ = (*β*_0_ *+ β*_1_) /2) The seasonal forcing on the transmission rate is considered (*E* = 1).
- **Scenario 3**: The restrictions are completely lifted after *t*_*int*_ (*β*_2_ =*β*_0_). The seasonal forcing on the transmission rate is not considered (*E* = 0).
- **Scenario 4**: The restrictions are partially lifted after *t*_*int*_ (*β*_2_ = (*β*_0_ + *β*_1_) /2). The seasonal forcing on the transmission rate is not considered (*E* = 0).

### 2.2 Parameter fitting and numerical solutions

In order to enhance the stability of the traditional least square method (Gauss-Newton algorithm), we use an improved damped least square method called Levenberg-Marquardt algorithm^17^. This method inserts a damping coefficient into the Gauss-Newton method when calculating the Hessian matrix. The benefit of introducing this damping coefficient is that it can converge very quickly in the steepest direction in many cases even when the initial solution is very far from ideal values, which makes the parameter determination more robust^18^. In addition, for all damping coefficient greater than 0, the coefficient matrix is positive definite which makes the Hessian matrix in the descending direction. The input variables to obtain fitted parameters(*α, β, γ, δ, λ*, and *κ*) are the time series of confirmed (*Q* (*t*) − *D* (*t*) − (*R* (*t*)), death,(*D* (*t*)), and recovered (*R* (*t*)) cases provided by from Johns Hopkins University Center for Systems Science and Engineering. The equations are solved using the classic 4th order Runge-Kutta method.

## 3 Projections of the US cities

Unfortunately, the recent protests against police violence in cities across the United States have gone ahead despite the current rising COVID-19 pandemic and potential subsequent outbreaks, possibly with higher intensity. Large public gatherings, shouting, and marching shoulder to shoulder may have already sown the seeds of the second outbreaks in regions under initial control^19,20^ and made it even more difficult to contain the epidemic in regions where the curve is still increasing. The use of tear gas and pepper spray against the protesters may also have produced violent coughing and runny noses, forcing protesters to remove their masks and making the crowds even more susceptible to the virus. A certain number of patients with the latent disease may have participated in the protests and spread the disease to healthy protesters, police officers, and national guards who are not yet immune to the virus^21^. If the close contacts of the infectious are not fully tracked, they may spread the virus to other groups of people, increasing the risk of a larger size of outbreaks. Here, we simulated the impact of large-scale protests on the potential second outbreaks in several cities of the United States^22^ (Figure 2 and Table 1). The model generally predicted a second wave of COVID-19 in the second half of 2020. We estimated the increase in the population of potentially infected people (*δ*E_t_) for each city based on the ratio of the number of infected persons (Q_t_) to the total population of the city (N). The timing of protests and the number of protesters in each city were collected from local news reports (Table 1). The increase in the population of potentially infected people (*δ*E_t_) and the populations of protesters (*δ*S_t_, regarded as an increase in the population of susceptible) were used as the force input for the model calculations to simulate the impact of protests on the outbreaks. When the protests begin, we force group E and group S to increase *δ*E_t_ and *δ*S_t_, respectively.

**Table 1.** Projections of the second outbreaks in some US cities in Scenario 1.

| Countries | Population<br>(million) | Start date of<br>Protests | Estimated<br>participants | The peak time<br>of the second<br>outbreaks | Peak time daily new<br>incidence without protests | Peak time daily new<br>incidence with protests | End of the second<br>outbreak | Accumulated<br>Confirmed Cases |
| --- | --- | --- | --- | --- | --- | --- | --- | --- |
| New York City (New York) | 8.33 | May, 29<sup>th</sup> | 25,000 | Mid-August | Around 8,000 | Around 18,000 | End of 2020 | Around 900,000 |
| Chicago (Illinois) | 5.15 | May, 28<sup>th</sup> | 30,000 | Mid-September | Around 2700 | Around 4100 | End of 2020 | Around 390,000 |
| Minneapolis (Minnesota) | 1.26 | May, 26<sup>th</sup> | 30,000 | Mid-September | Around 400 | Around 700 | End of 2020 | Around 48,000 |
| Columbus (Ohio) | 1.31 | May, 28<sup>th</sup> | 10,000 | Late-September | Around 120 | Around 380 | End of 2020 | Around 35,000 |
| Westchester (Illinois) | 0.97 | May, 29<sup>th</sup> | 30,000 | Mid-August | Around 3,500 | Around 5,500 | September, 2020 | Around 150,000 |
| Philadelphia<br>(Pennsylvania) | 10.04 | May, 30<sup>th</sup> | 10,000 | Mid-October | Around 800 | Around 1,700 | End of 2020 | Around 120,000 |
| Seattle (Washington) | 2.25 | May, 29<sup>th</sup> | 50,000 | Late-August | Around 2,00 | Around 600 | End of 2020 | Around 35,000 |
| Washington, D.C. | 0.70 | May, 29<sup>th</sup> | 10,000 | Mid-October | Around 180 | Around 650 | September, 2020 | Around 70,000 |
| San Francisco (California) | 0.88 | May, 30<sup>th</sup> | 30,000 | Mid-August | Around 100 | Around 400 | October, 2020 | Around 22,000 |
| Detroit (Michigan) | 1.75 | May, 29<sup>th</sup> | 30,000 | Early-September | Around 1,000 | Around 2,200 | October, 2020 | Around 70,000 |
| Miami-Dade (Florida) | 2.72 | May, 30<sup>th</sup> | 20,000 | Mid-August | Around 1,800 | Around 3,000 | September, 2020 | Around 120,000 |
| San Diego (California) | 3.34 | May, 29<sup>th</sup> | 30,000 | Late-August | Around 1,000 | Around 1,600 | September, 2020 | Around 130,000 |

**Figure 2:**
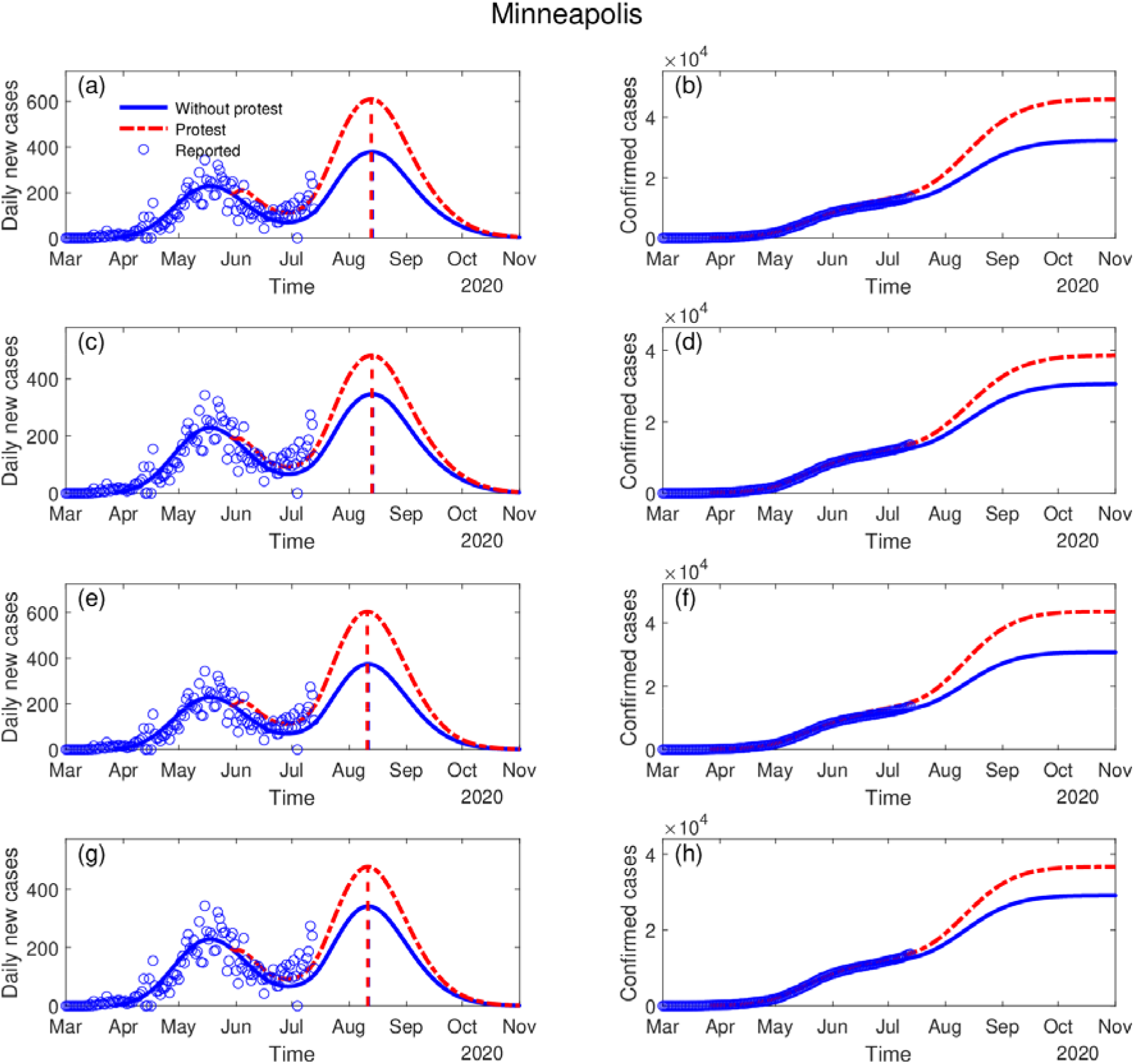
The impact of protests on the possible second outbreak in Minneapolis. The blue dots denote the reported daily incidence of COVID-19 cases. The blue line represents the simulation and projections without protests, while the dashed red line denotes the simulation and projections with protests. Four scenarios with protests and four scenarios without protests are simulated. (a)∼ (b), (c)∼(d), (e)∼(f), (g)∼(h) are the simulation for Scenario 1, 2, 3, 4, respectively. (a)∼(b) and (c)∼(d) are the simulations with seasonal forcing; (e)∼(f) and (g)∼(h) are the simulations without seasonal forcing. (a)∼(b) and (e)∼(f) are the simulations where the restrictions are completely lifted; (c)∼(d) and (g)∼(h) are the simulations where the restrictions are partially lifted.

Figure 2 shows the projections for Minneapolis in 8 scenarios (4 scenarios with protests and 4 without). After the COVID-19 restrictions were lifted, an upward trend of daily new cases has been observed. The protests would significantly amplify the intensity of the second outbreak but may not be able to advance it. In scenarios 1, the second outbreak of Minneapolis will peak in mid-August 2020. The comparison between scenarios indicates that the effect of intervention measures outweighs the seasonal forcing. For the rest of the 12 cities, the model also predicted enhanced second outbreaks when the impact of protests is considered (Table 1). Due to space constraints, the details of the projection results are not presented in the manuscript and can be accessed at http://covid-19.lzu.edu.cn/.

## 4 Projections of countries with reported protests

In addition to the United States, protests of a certain scale also broke out in other countries. Using similar parameterization of the protests, Table 3 presents the projections of the second outbreaks in the United Kingdom, the United States, Germany, Italy, Australia, Canada, Spain, Mexico, Switzerland, Belgium, Netherlands, Ireland, and Denmark. For the United Kingdom (Figure 3), the second outbreak is likely to peak during August. Under the impact of protests, when the restrictions are lifted completely, a second wave with a peak of 14,160 is expected, which is 75.6% higher than the scenario without protests (Scenario 1). The protests and the lifting of restrictions, along with the enhancement in the ability of virus transmission in the cold seasons due to temperature change may cause the recurrence of an outbreak that was initially under control. If the same intervention measures are implemented during the second outbreak, the second outbreak would be brought under control again by the end of 2020.

**Table 2.** Projections of the second outbreaks in some countries (with protests)

| Countries | Population<br>(million) | Start date<br>of Protests | Estimated<br>participants | The peak time<br>of the second<br>outbreaks | Peak time daily new<br>incidence without<br>protests | Peak time daily new<br>incidence with<br>protests | End of the second<br>outbreak | Accumulated<br>Confirmed Cases<br>with protests |
| --- | --- | --- | --- | --- | --- | --- | --- | --- |
| United Kingdom | 66.48 | May, 28<sup>th</sup> | 70,000 | Early-August | Around 8,000 | Around 14,170 | End of 2020 | Around 1,150,000 |
| United States | 328.2 | May, 26<sup>th</sup> | 1,500,000 | Late-July | Around 40,000 | Around 120,000 | End of 2020 | Around 16,000,000 |
| Germany | 82.93 | May 30<sup>th</sup> | 160,000 | Early-September | Around 4,500 | Around 7,000 | End of 2020 | Around 450,000 |
| Italy | 60.43 | June, 30<sup>th</sup> | 150,000 | Early-October | Around 6,000 | Around 95,000 | End of 2020 | Around 600,000 |
| Australia | 25.44 | June, 2<sup>nd</sup> | 14,000 | Early-August | Around 500 | Around 1,900 | End of 2020 | Around 45,000 |
| Canada | 37.05 | May, 30<sup>th</sup> | 100,000 | Late-September | Around 1,800 | Around 2,500 | End of 2020 | Around 250,000 |
| Spain | 46.73 | June, 1<sup>st</sup> | 10,000 | Early-September | Around 7,600 | Around 9,400 | End of 2020 | Around 1,100,000 |
| Mexico | 126.2 | May, 30<sup>th</sup> | 50,000 | Late-August | Around 5,800 | Around 7,600 | End of 2021 | Around 1,500,000 |
| Switzerland | 8.57 | May, 31<sup>st</sup> | 20,000 | Early-August | Around 1,400 | Around 2,100 | October, 2020 | Around 80,000 |
| Belgium | 11.42 | June, 1<sup>st</sup> | 50,000 | Late-August | Around 2,000 | Around 4,200 | End of 2020 | Around 230,000 |
| Netherlands | 17.26 | June, 1<sup>st</sup> | 56,000 | Late-July | Around 1,800 | Around 2,800 | End of 2020 | Around 170,000 |
| Ireland | 4.81 | May, 31<sup>st</sup> | 20,000 | Mid-July | Around 600 | Around 1,500 | End of 2020 | Around 90,000 |
| Denmark | 5.73 | May 31<sup>st</sup> | 20,000 | Late-August | Around 600 | Around 1,000 | End of 2020 | Around 60,000 |

**Table 3.** Projections of the second outbreaks in some countries in Scenario 1 (without reported protests)

| Countries | Population<br>(million) | The peak time of the second<br>outbreaks | Peak time daily new incidence without<br>protests | End of the second<br>outbreak | Accumulated Confirmed<br>Cases |
| --- | --- | --- | --- | --- | --- |
| India | 1324 | Early-November, 2020 | Around 41,000 | February, 2021 | Around 6,000,000 |
| Russia | 144.5 | Late-September, 2020 | Around 12,000 | April, 2021 | Around 2,800,000 |
| Brazil | 209.5 | Early-October, 2020 | Around 60,000 | February, 2021 | Around 11,200,000 |
| Peru | 32.05 | Late-October, 2020 | Around 13,000 | May, 2021 | Around 2,500,000 |
| Chile | 18.60 | Late-July, 2020 | Around 11,000 | April, 2021 | Around 1,500,000 |
| Argentina | 44.49 | January, 2021 | Around 5000 | April, 2021 | Around 600,000 |
| Pakistan | 212.2 | Early-August, 2020 | Around 30,000 | November, 2020 | Around 2,700,000 |
| Saudi<br>Arabia | 32.55 | Late-July, 2020 | Around 13,000 | May, 2021 | Around 3,500,000 |
| Bangladesh | 161.4 | Late-August, 2020 | Around 15,000 | January, 2021 | Around 1,600,000 |
| Qatar | 2.64 | Early-September, 2020 | Around 2,400 | December, 2020 | Around 240,000 |
| Colombia | 49.64 | February, 2021 | Around 12,500 | May, 2021 | Around 55,000 |
| Belarus | 9.51 | Mid-August, 2020 | Around 2,800 | May, 2021 | Around 2,800,000 |
| Egypt | 102.27 | Mid-November, 2020 | Around 2,800 | March, 2021 | Around 420,000 |
| Ecuador | 17.08 | Mid-August, 2020 | Around 5,000 | December, 2020 | Around 300,000 |

**Figure 3:**
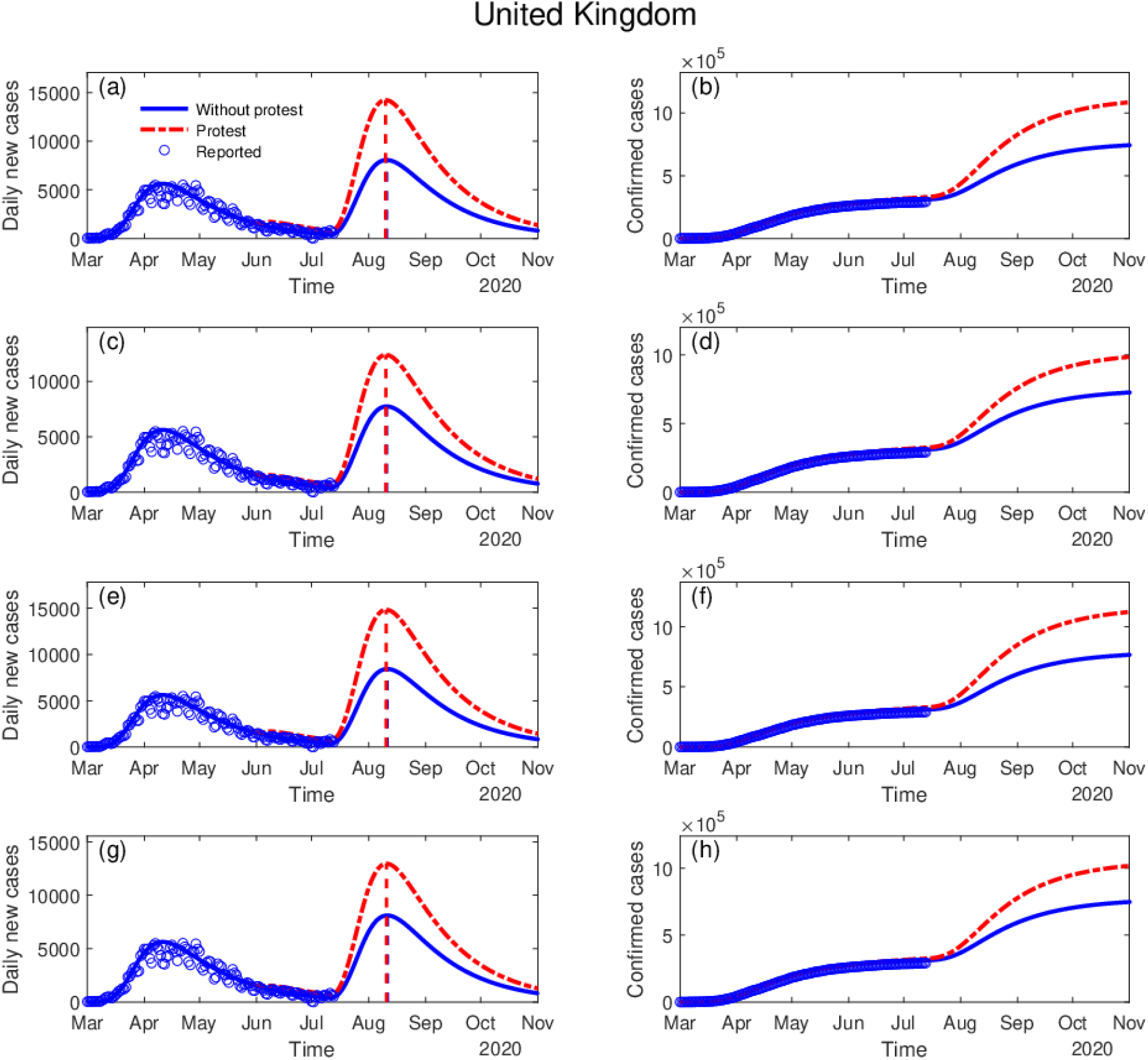
The impact of protests on the possible second outbreak in the United Kingdom. The blue dots denote the reported daily incidence of COVID-19 cases. The blue line represents the simulation and projections without protests, while the dashed red line denotes the simulation and projections with protests. The scenarios in these subplots are the same as Figure 2.

## 5 Projections of countries without reported protest

Mass gathering events during the epidemic should be restricted or even banned since they have the potential to enhance the second outbreak and pose further radical public-health challenges for health authorities and governments^23,24^. Lifting the restrictions too early may also have the potential to trigger subsequent outbreaks and further increase the pressure on the medical system. Fig 4 shows the projections of India in the four scenarios. With seasonal forcing, it is predicted that the first peak of the epidemic will occur in September while the second peak with higher intensity, caused environmental changes, will arrive in January 2021. Without seasonal forcing, there would be only one peak in September 2020. We also projected the epidemic curve for other countries including Russia, Brazil, Chile, etc. that are still in the rapid growing period. We classified them as ‘non-protesting countries’, not because there are no protests. Indeed, there might have been many protests and mass gatherings in India, Brazil, and other regions that may impact the outbreak on varying degrees, but the information on the timing and size of these protests are currently not available. Therefore, the role of these protests on the timing and size of the outbreaks can not be isolated and may not be incorporated as a force into the model.

**Figure 4:**
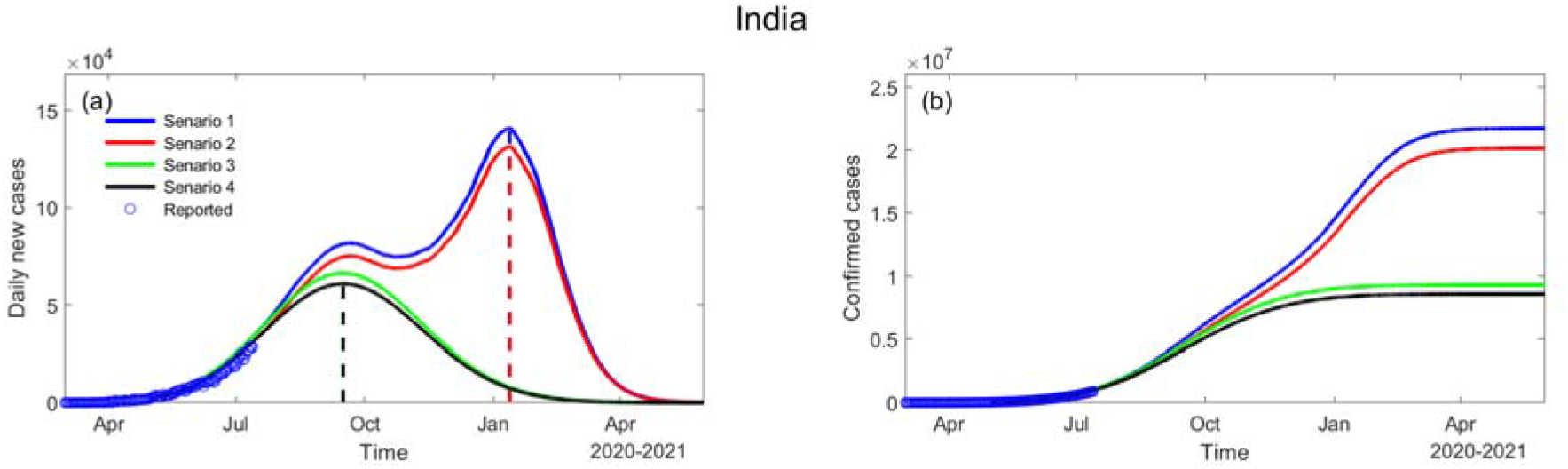
Projections of the second outbreak in India in different scenarios. Scenarios 1 and 2 are simulated with seasonal forcing, while scenarios 3 and 4 are simulated without seasonal forcing. Scenarios 1 and 3 are the projections where restrictions are fully lifted, while scenarios 2 and 4 are the projections where restrictions are partially lifted.

From Table 3 we can see that the peak time and size of the second outbreak varies from countries to countries, due to different levels of interventions measures, environmental conditions, medical resources, etc. Regions with high population density are at higher risk of enhanced second outbreaks. When control measures are lifted too soon and environmental temperatures are more suitable for the spread of disease^25^, an enhanced second outbreak is expected. Therefore, when considering the timing of lifting the restrictions to restore the economy, it is necessary to analyze epidemic situations as well as climate factors. For example, during cold seasons when the transmission rate is higher, reopening would easily light up the second outbreaks, since conducive environmental factors and related human social behaviors (more frequent indoor gatherings) would, directly and indirectly, increase the transmission ability of the virus, leaving more people vulnerable to infection. Therefore, the peak of the second wave of the outbreak is most likely to synchronize with the fall of environmental temperature, displaying strong seasonality. During winter months, the temperate regions of the Northern and Southern Hemispheres experience highly synchronized annual influenza epidemics^26^. Additionally, when restrictions are lifted, personal protection (wearing face masks, keeping appropriate interpersonal distance, sterilization, etc)^27^. is still required or even mandatory in indoor places so as to minimize the infection rate by cutting off the infection routine.

## 6 Discussion and conclusion

New treatments and vaccines are not yet available for any COVID-19-affected areas^28^. With the presence asymptomatic of carriers that may spread the virus, and the lack of herd immunity, a second outbreak is inevitable as confirmed cases of COVID-19 increase. Our results show that the timing and intensity of the second outbreaks are seasonally modulated and depend largely on local reopening policies. Higher seasonal variations in COVID-19 transmission may lead to a greater incidence of recurrent wintertime outbreaks^29^. The current mass protests in the United States and other regions of the world could lead to amplified second outbreaks, overlapping the wintertime outbreaks and threatening more lives. This is because the extremely crowded environments facilitate the spread of the virus, leading to high rates of the second attack, as seen in both the 1918 pandemic and the 1957 Asian influenza pandemic^30–32^.

If the transmission capacity of the second outbreak increases, it could place a catastrophic burden on the health system and create even more serious social and economic crises. However, if the chain of transmission is cut during the first outbreak, there will be no further outbreak similar to the first wave. The necessary drug therapies and vaccines currently require long-term development and testing, so nonpharmaceutical interventions are the only direct means available to suppress the spread of the disease^33^. In the face of a powerful pandemic, everyone must help to fight the virus. We must collectively follow the distancing limits recommended by global public-health organizations to effectively reduce the potential cost in the lives of the second wave of infection. Our findings should encourage close monitoring and early warning of the development of a global pandemic, as well as safeguards for our global prediction system to best contain the second wave^34^. These results provide a powerful scientific basis for governments to adjust their policies and control measures in real-time, to achieve the most effective allocation of medical resources before the second outbreak and to reduce the associated health risks.

## Data Availability

The data in this paper are publicly available at http://covid-19.lzu.edu.cn/.

## Acknowledgments

The authors acknowledge the Center for Systems Science and Engineering (CSSE) at Johns Hopkins University for providing the COVID-19 data. We acknowledge E. Cheynet for providing the Generalized SEIR Epidemic Model (fitting and computation). This work was jointly supported by the National Science Foundation of China (41521004) and the Gansu Provincial Special Fund Project for Guiding Scientific and Technological Innovation and Development (Grant No. 2019ZX-06).

